# Unusual predominance of *Staphylococcus aureus* in the salivary microbiome of children with Early Childhood Caries in Kano, Nigeria

**DOI:** 10.64898/2026.03.05.26347684

**Authors:** C.C Okolo, T.G Amole

## Abstract

**Background:** The microbial aetiology of early childhood caries (ECC) in sub-Saharan African populations remains poorly characterised, with most studies focusing on conventional cariogenic pathogens like Streptococcus mutans. This study aimed to characterise the salivary microbial profile of children with ECC in urban Kano, northern Nigeria.

**Methods:** In this cross-sectional study of 162 children aged 3–5 years in urban Kano, unstimulated saliva samples were collected and analysed using standard bacteriological culture methods. Caries status was assessed using decayed, missing, and filled teeth (dmft) index and International Caries Detection and Assessment System (ICDAS). Microbial isolates were identified through Gram staining, colony morphology, and biochemical tests (catalase, coagulase, oxidase).

**Results:** Of 32 microbial isolates obtained, Staphylococcus aureus was the most prevalent (43.8%, n=14), followed by Streptococcus species (28.1%, n=9), Klebsiella species (12.5%, n=4), non-aureus staphylococci (6.3%, n=2), yeast (6.3%, n=2), and Pseudomonas species (3.1%, n=1). Only one isolate demonstrated direct association with dmft-detectable caries. Polymicrobial colonisation occurred in four cases (12.5%), predominantly featuring S. aureus-yeast combinations (n=2). White spot lesions (ICDAS 1-2) were associated with S. aureus and Klebsiella species in two separate cases.

**Conclusion:** This study reveals an unexpected predominance of S. aureus in the salivary microbiome of children in northern Nigeria, challenging conventional paradigms of ECC microbiology. The low correlation between microbial isolates and clinical caries suggests complex, multifactorial aetiology. These findings highlight the need for molecular characterisation of oral microbiomes in African populations and reconsideration of caries pathogenesis models in this unique epidemiological context.

## Introduction

Early childhood caries (ECC) remains a significant global public health concern, affecting approximately 48% of preschool children worldwide [1]. Traditional understanding of caries pathogenesis emphasises the central role of specific acidogenic and aciduric bacteria, particularly *Streptococcus mutans* and *Lactobacillus* species, which metabolise dietary sugars to produce organic acids that demineralise tooth enamel [2]. However, contemporary molecular approaches have revolutionised our understanding of caries microbiology, revealing complex polymicrobial ecosystems and ecological shifts rather than simple pathogen colonisation [3].

The ecological plaque hypothesis posits that caries develops through disruption of microbial homeostasis, with environmental factors (particularly frequent sugar exposure) selecting for acid-tolerant communities capable of surviving and thriving in low-pH conditions [4]. While *mutans streptococci* remain important keystone pathogens, numerous other bacteria including *Scardovia wiggsiae, Actinomyces* species, *Bifidobacterium* species, and the fungus *Candida albicans* have been implicated in caries pathogenesis through various metabolic pathways and synergistic interactions [5,6].

Despite advances in understanding oral microbiome dynamics in caries development, significant geographical gaps persist in microbial characterisation, particularly across sub-Saharan Africa. Nigerian data on the microbiology of ECC remain sparse: only a single published investigation exists, and it employed culture-dependent methods focused exclusively on *Streptococcus mutans* in a southern Nigerian population [7]. This is a notable limitation, given the well-established influence of environmental and host-related factors — including diet, endogenous nutrient availability, immune function, systemic disease, and antimicrobial exposure — on oral microbial community composition [8,9]. Northern Nigeria presents a particularly distinct ecological context, characterised by an arid Sahelian climate, traditional dietary patterns, culturally specific oral hygiene practices, widespread non-prescription antibiotic use, and a predominantly Muslim population whose religious practices may further shape oral health behaviours. To date, no study has characterised the oral microbiome of children with ECC in this setting, representing a critical gap in both the regional and global literature.

*Staphylococcus aureus*, traditionally considered an opportunistic pathogen associated with skin and soft tissue infections, has recently gained attention as a potential oral commensal with emerging roles in oral diseases [10]. Classified by the World Health Organization as a High Priority Pathogen due to antimicrobial resistance concerns, *S. aureus* has been detected in various oral niches, though its significance in caries pathogenesis remains controversial [11,12]. Some studies suggest synergistic interactions between *S. aureus* and cariogenic bacteria, while others propose competitive exclusion mechanisms [13].

This study aimed to characterise the salivary microbial profile of children with ECC in urban Kano, northern Nigeria, and explore associations between specific microorganisms and caries presence and severity. Given the unique sociocultural and environmental context of northern Nigeria, we hypothesised that the oral microbiome of children in this region might differ from patterns reported in other populations.

## Methods

### Study Design and Setting

This analytical cross-sectional study was conducted as part of a larger investigation of ECC prevalence and determinants in urban Kano, northwest Nigeria, between 30^th^ of September and 4^th^ of November 2024. The study received ethical approval from the Kano State Health Research Ethics Committee (NHREC/17/03/2018).

### Study Population

The study included 162 children aged 36–71 months recruited through multistage sampling from Kano Municipal and Nasarawa Local Government Areas. Inclusion criteria required children to have teeth present for clinical assessment, parental informed consent, and cooperation for oral examination. Children with acute oral infections or debilitating conditions were excluded.

### Clinical Examination and Caries Assessment

Clinical oral examinations were performed by calibrated dental surgeons using sterile mouth mirrors and WHO periodontal probes under artificial illumination. Caries diagnosis employed two systems: the traditional decayed, missing, and filled teeth (dmft) index and the International Caries Detection and Assessment System (ICDAS). ICDAS codes 1-2 represented early disease (white spot lesions), codes 3-4 moderate disease, and codes 5-6 advanced disease.

### Salivary Sample Collection

Unstimulated whole saliva samples (approximately 2 mL) were collected from each participant using the passive drool method. Children refrained from eating, drinking, or oral hygiene procedures for at least 30 minutes before collection. Samples were obtained in sterile 15 mL centrifuge tubes, immediately placed in cooler boxes with ice packs (2-8°C), and transported to the Africa Centre for Excellence in Population Health and Policy (ACEPHAP) Molecular Laboratory within four hours.

### Microbiological Processing

Laboratory analysis followed standard bacteriological protocols under biosafety level 2 conditions. Each saliva sample was vortexed for homogenisation, and 10 µL aliquots were streaked onto three culture media: blood agar (5% sheep blood), MacConkey agar, and chocolate agar. Plates were incubated aerobically at 37°C for 24-48 hours, with blood and chocolate agar plates additionally incubated in 5-10% CO_2_.

### Microbial Identification

Colony morphology assessment included size, shape, elevation, margin, surface characteristics, opacity, colour, and haemolysis patterns. Gram staining distinguished Gram-positive from Gram-negative organisms. Biochemical identification included:

a. Catalase test (3% hydrogen peroxide) to differentiate staphylococci (positive) from streptococci (negative)
b. Coagulase test (rabbit plasma) to identify S. aureus
c. Oxidase test for Gram-negative isolates
d. Additional biochemical characterisation as needed

Organisms were categorised as: *Staphylococcus aureus, Streptococcus* species, *Klebsiella* species, non-aureus staphylococci, yeast/fungi, and other bacteria. Polymicrobial colonisation was defined as isolation of two or more distinct microbial species from a single sample.

### Quality Control

To ensure reliability, all culture media were subjected to sterility testing and performance verification using reference strains obtained from the American Type Culture Collection (ATCC): *Staphylococcus aureus* ATCC 25923, *Escherichia coli* ATCC 25922, and *Streptococcus pneumoniae* ATCC 49619. Biochemical reagents used for microbial identification were validated in each round of testing by including appropriate positive and negative controls. Furthermore, to assess the reproducibility of the identification process, a random subset of isolates (10%) was independently re-identified by a second, blinded laboratory scientist; concordance was observed for all isolates tested.

### Data Analysis

Data were analysed using IBM SPSS Statistics version 26.0. Descriptive statistics characterised microbial distribution. Chi-square or Fisher’s exact tests examined associations between specific microorganisms and caries presence/severity. Given the exploratory nature of this microbiological investigation and limited positive cultures relative to sample size, multivariate analysis was not performed for microbial variables.

## Results

### Microbial Isolation Patterns

Of 162 saliva samples processed, 32 microbial isolates were obtained from 28 children (17.3% of participants). *Staphylococcus aureus* was the most prevalent organism, followed by *Streptococcus* species, and *Klebsiella* species.

**Table 1:**
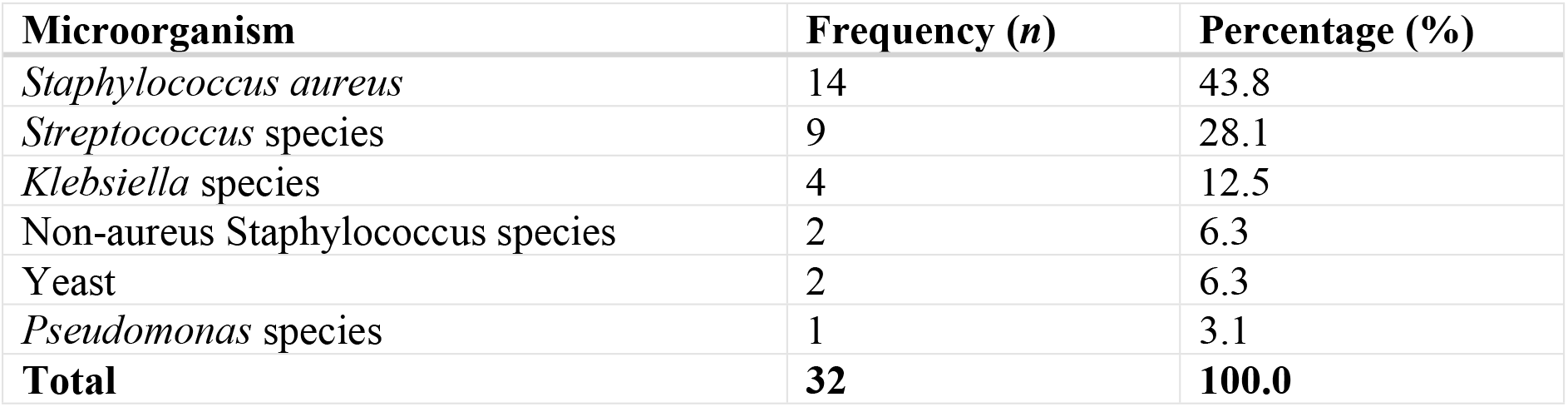
Distribution of microbial isolates from salivary samples (n=32)

### Polymicrobial Colonisation

Polymicrobial colonisation occurred in four cases: the most frequent combination was *S. aureus* with yeast (*n*=2), followed by *S. aureus* with *Streptococcus* species (*n*=1), and one case of triple colonisation with *S. aureus, Streptococcus species*, and *Klebsiella* species (*n*=1) (Table 2).

**Table 2:**
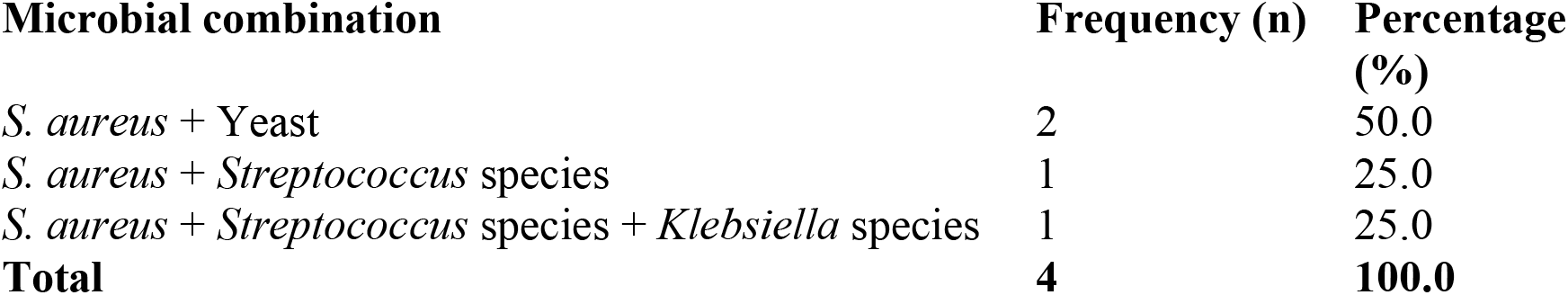
Patterns of polymicrobial colonisation (*n*=4)

### Association with Caries Status

Only one of the 32 microbial isolates (3.1%) demonstrated direct association with dmft-detectable caries (dmft >0), identified as a *Streptococcus* species (Table 3). The remaining 31 isolates (96.9%) came from children with dmft = 0, despite positive microbial cultures.(Table 3). When examining early caries lesions (ICDAS 1-2, white spot lesions), two samples from affected children yielded microbial growth: one with *S. aureus* and another with *Klebsiella* species (Table 3).

**Table 3:** Summary of microbial isolates by caries status.

| Caries status | Microbial species | Number of isolates | Percentage (%) |
| --- | --- | --- | --- |
| dmft = 0 | Various (non-Streptococcus) | 31 | 96.9 |
| dmft > 0 | *Streptococcus* species | 1 | 3.1 |
| White spot (ICDAS 1-2) | *Staphylococcus aureus* | 1 | 50%* |
| White spot (ICDAS 1-2) | *Klebsiella* species | 1 | 50%* |

\*Percentage of white spot lesion samples with microbial growth (n=2)

## Discussion

This study reveals an unexpected salivary microbial profile among preschool children in northern Nigeria, characterised by the predominance of *Staphylococcus aureus* rather than the mutans streptococci that conventional frameworks position as the primary aetiological agents of early childhood caries (ECC). This finding challenges well-established paradigms in caries microbiology and raises important questions about the extent to which globally derived microbiological models can be applied to geographically and ecologically distinct populations.

The microbial profile observed in this study contrasts from that reported in the majority of ECC investigations worldwide, where *S. mutans* consistently emerges as the predominant cariogenic isolate and a robust predictor of disease[2,7]. Systematic reviews and microbiome-based meta-analyses have confirmed that *S. mutans* prevalence and relative abundance are reliably elevated in caries-active compared to caries-free children, with its presence associated with approximately four-fold higher odds of ECC development, and specific clades linked to up to 33-fold increased risk in some cohort studies[14]. The contrast is rendered particularly striking by comparison with the only analogous Nigerian study — Iyun et al., conducted in Ibadan in southern Nigeria — in which *S. mutans* was detected in all caries-affected and caries-free children examined[7]. The microbiological picture in that southern cohort thus differs markedly from the one described here.

A growing body of evidence demonstrates that oral microbiome composition varies substantially across geographies and populations, shaped by differences in diet, climate, oral hygiene practices, socioeconomic conditions, and host genetic background[15,16]. While a conserved core salivary microbiome exists across human populations, regional shifts in non-core taxa and disease-associated community profiles are well-documented, and African as well as other non-industrialised populations frequently exhibit distinct plaque and salivary communities[15]. Several molecular studies have also reported *S. mutans* at low abundance or entirely undetectable in some caries lesions, reinforcing a polymicrobial, dysbiosis-based model of disease in which no single species is universally obligate[3,5,6]. The present findings contribute to this expanding evidence base and suggest that global ECC microbiology is considerably more heterogeneous than classical frameworks have assumed[3].

Several contextual factors unique to northern Nigeria may collectively account for the striking predominance of *S. aureus* in this population. Widespread, often unsupervised antibiotic use likely disrupts the competitive dominance that commensal streptococci normally maintain within the oral cavity, creating ecological niches amenable to opportunistic, antimicrobial-tolerant organisms such as *S. aureus*[17,18]. This mechanistic reasoning is supported by analogous observations from gut microbiome research and other Nigerian clinical settings, though direct oral microbiome evidence from this region remains absent, and the proposed mechanism should be understood as biologically and epidemiologically plausible rather than confirmed [17,18]. Concurrently, the region’s well-documented consumption of raw and traditionally fermented dairy products — including *nono, kindirmo*, and *fura-da-nono* — represents a plausible reservoir and transmission route for *S. aureus*, with multiple Nigerian studies documenting staphylococcal prevalence of 5–12% in these food sources, alongside frequent multidrug-resistant phenotypes [19-21].

Two further environmental and behavioural factors may reinforce this ecological shift. The arid Sahelian climate, characterised by extreme thermal variation and persistently low humidity, plausibly selects for stress-resilient microorganisms; broader modelling and experimental data support temperature-linked advantages in *S. aureus* survival and habitat suitability, though oral-site evidence from Sahelian populations has yet to be generated [22,23]. Additionally, the habitual use of *miswak* (*Salvadora persica*) as a traditional oral hygiene implement — while broadly beneficial — appears to exert substantially stronger antimicrobial activity against streptococci than against *S. aureus* in both in vitro and clinical studies, potentially tilting the oral microbial balance in favour of staphylococcal persistence [24,25].

The clinical significance of oral *S. aureus* colonisation extends well beyond its putative contribution to caries pathogenesis. The oral cavity is increasingly recognised as an independent reservoir for antimicrobial-resistant *S. aureus* strains, with carriage rates broadly comparable to those reported for nasal colonisation in healthy individuals, and oral isolates frequently demonstrating multidrug resistance [11,12]. Oral *S. aureus* colonisation in young children is therefore clinically meaningful in the context of antimicrobial resistance surveillance, as it plausibly represents a source for systemic dissemination following mucosal breach — for instance, during dental procedures — paralleling established evidence linking oral bacteraemia to infective endocarditis and other systemic sequelae. Given *S. aureus*’s designation as a WHO High Priority Pathogen on account of antimicrobial resistance concerns, these implications warrant heightened attention in paediatric dental infection control protocols serving this population[11,12].

The weak association observed between microbial isolation and clinically detectable caries in this cohort is consistent with the contemporary ecological model of caries, which frames disease not as the product of single-pathogen infection but as the outcome of biofilm-level dysbiosis driven by diet, host factors, and microbial community dynamics [3]. Established evidence indicates that both putatively cariogenic and non-cariogenic taxa are routinely recovered from healthy and diseased oral sites alike; what distinguishes disease from health is not pathogen presence per se, but an acidogenic, low-diversity community shift arising from ecological disruption. This is further reinforced by the polymicrobial patterns documented in this cohort [3].

This study has several limitations that should be considered when interpreting its findings. The culture-dependent approach employed, whilst standard in clinical microbiology practice, inevitably underestimates microbial diversity by selecting for readily cultivable organisms; many oral bacteria are fastidious or require specific growth conditions not afforded by the media used here [3]. Molecular methods such as 16S rRNA gene sequencing or metagenomic approaches would provide substantially more comprehensive community profiling, but were beyond the resources available for this study[26]. Furthermore, the cross-sectional design precludes determination of whether the observed microbial patterns preceded caries development or arose as a consequence of ecological changes following disease onset. Although standardised pre-collection instructions were provided to minimise variability, sample collection timing relative to meals, oral hygiene activities, or recent antibiotic use may have influenced the results to some degree.

## Conclusion

This study documents a distinctive salivary microbial profile in preschool children in northern Nigeria, marked by the unexpected predominance of *Staphylococcus aureus* over conventional cariogenic streptococci. The weak correspondence between microbial isolation and clinical caries reinforces the multifactorial, ecological nature of ECC pathogenesis and cautions against single-pathogen interpretive frameworks. These findings challenge prevailing caries microbiology paradigms and underscore the necessity of context-sensitive microbiological characterisation in diverse global populations. Molecular characterisation of oral microbiomes across Nigerian geographical regions is urgently warranted, alongside longitudinal investigation of the temporal relationships between microbial colonisation patterns and caries development, and systematic profiling of antimicrobial resistance among oral *S. aureus* isolates in this setting.

## Data Availability

All relevant data are within the manuscript and its Supporting Information files.

## Acknowledgements

The authors are grateful to the children and their parents or guardians who consented to participate in this study. We thank the staff of the ACEPHAP Molecular Laboratory, Bayero University, Kano, for their technical support, and our research assistants for their diligence throughout data collection. We also acknowledge Professor Isa Abubakar for his oversight of laboratory activities.

## Data Availability

The microbial data supporting this study are available from the corresponding author upon reasonable request.

## Ethics Approval

Approved by Kano State Health Research Ethics Committee (NHREC/17/03/2018).

## Authors’ Contributions

CCO: Conceptualisation, methodology, investigation, writing – original draft.

TGA: Supervision, writing – review & editing.

All authors approved the final manuscript.

